# Occupational profile and prevalence of workplace accidents among beach workers

**DOI:** 10.1101/2025.01.15.25320620

**Authors:** Cleber Cremonese, Adedayo Michael Awoniyi, Mariela Sousa dos Santos, Jailma dos Santos Silva, Thayane Silva Nunes, Wiler de Paula Dias, Joelma Marques Rodrigues, Juliana Cristina dos Santos Soares, Armando Meyer

## Abstract

**Background:** Workplace accidents (WAs) are acute, often preventable events that result in injuries or functional impairments, typically arising from occupation-related activities. In 2019, an estimated 395 million workers worldwide suffered non-fatal work-related injuries, with 330,000 fatalities, the majority occurring in low- and middle-income countries (LMICs) like Brazil. The true figures may be considerably higher due to underreporting, as many informal workers lack adequate social protection and may avoid reporting accidents for fear of jeopardizing their livelihood.

**Objective:** To assess the prevalence of WAs and their association with demographic and occupational determinants among beach workers in Salvador, Bahia, Brazil, between 2023 and 2024.

**Methods:** A cross-sectional epidemiological study was conducted with 579 urban beach workers in Salvador from November 2023 to March 2024. Following ethical approval and participants’ consent, questionnaires were administered to gather data on sociodemographic and occupational characteristics, workplace environments and processes, workplace-related injuries and illnesses, and history and characteristics of WA. WA prevalence and prevalence ratios were calculated, and associations were analyzed using a Poisson regression model with robust variance.

**Results:** Among the workers, 59.4% were males, 25% were aged ≤29 years and 11.4% were ≥60 years. Black and brown individuals represented 92.9% of the population. The most performed activities were street vendors (43.6%) and waiters (25%). Informal employment was reported by 72.3% of workers, 70.2% worked ≥9 hours per day, and 88% had no occupational training. The overall WA prevalence observed was 40.3%, with workers ≤29 years old having a 2.59 times higher likelihood of experiencing WA compared to those ≥60 years old. The most common WAs were punctures (42.7%), cuts (28.4%) and burns (12.1%).

**Conclusions:** The high prevalence of WA among beach workers, especially those ≤29 years old, may be associated with inadequate working conditions, including long working hours and lack of occupational training. These findings highlight the need for targeted interventions to improve working conditions and reduce WA risks.

## INTRODUCTION

Workplace accident (WA) refers to any acute event capable of causing bodily injury or functional impairment to a worker, arising from non-natural factors related to work activities(1). Such incidents may occur either during work-related activities and/or while commuting between home and workplace. Examples include cuts, falls from height, burns, punctures, electric shocks, road accidents and instances of interpersonal violence(1,2).

Every WA needs to be recognized as an unforeseen yet preventable event(3). Regardless of employment type, location or nature of activity, WAs primarily occur due to insufficient or ineffective collective protective measures, poor working conditions and a lack of adequate training and individual protective equipment provided by employers(4,5). Consequences of WAs can range from temporary to permanent loss or reduction of physical and mental capacities and in more severe cases fatality(1).

In 2019, a year before the COVID-19 pandemic, 395 million workers worldwide suffered non-fatal work-related injuries, resulting in 330,000 deaths, with the majority of cases occurring in low- and middle-income countries (LMICs) such as Brazil (6). In Brazil alone, 1,675,056 non-fatal WAs and 25,455 deaths were reported in the Notifiable Injury Information System (SINAN)-the primary national system for reporting accidents, diseases, and injuries, linked to the Unified Health System (SUS)-between 2014 and 2023(7). However, WAs in Brazil and globally are likely underreported due to the absence or precariousness of registration systems, and the limited training of professionals involved in monitoring and data collection. Similarly, the lack of social protection inherent to the informal nature of the participants’ work often discourages the reporting of WA, as workers may fear losing their jobs(8,9).

In July 2024, Brazil’s workforce reached 101 million, representing 46% of the population. Informal workers comprised 40% of this workforce, with higher representation among females, individuals with lower education levels, Black and Brown race, and residents in the North and Northeast regions of the country(10). With an extensive Atlantic coastline of approximately 7,000 kilometers, economic activities are prevalent along the Brazilian coast. In these areas, workers engage in various activities such as selling beverages, food, clothing, handicrafts and beauty products, as well as offering services like chair, umbrella and toy rentals. Other common occupations include surfing, swimming and diving instructors, waiters and lifeguards. Beach workers are often exposed to a range of workplace health risks, including prolonged sun exposure, contact with potentially contaminated sand and water, long hours traversing uneven terrain while carrying heavy loads, as well as potential territorial disputes, harassment and interpersonal violence(11,12).

Despite the presence of workers involved in economic activities on beaches in Brazil and other LMICs with similar characteristics, there remains a significant lack of knowledge regarding their demographic and occupational profiles, as well as their history of workplace injuries and illnesses. This knowledge gap, coupled with the absence of data from epidemiological studies and the apparent neglect by government institutions, contributes to the marginalization of these workers in terms of their work environments and processes, and health conditions. Therefore, this study aims to assess the prevalence of WAs in relation to demographic and occupational determinants among beach workers in Salvador, Bahia, Brazil between 2023 and 2024.

## METHODS

### Study design and area

A cross-sectional, observational epidemiological study was conducted among workers from five urban beaches in Salvador, the capital of Bahia, and the largest and most populous state in northeastern Brazil. Salvador, served as Brazil’s first capital (1549 to 1763), and has a current population of approximately 2.4 million(13). The sample included five beach areas with the highest concentration of workers, namely: Farol de Itapuã-Sereia, Armação-Piatã, Cristo-Porto da Barra, Boa Viagem-Ribeira and Tubarão-São Tomé de Paripe (Figure 1A).

**Figure 1:**
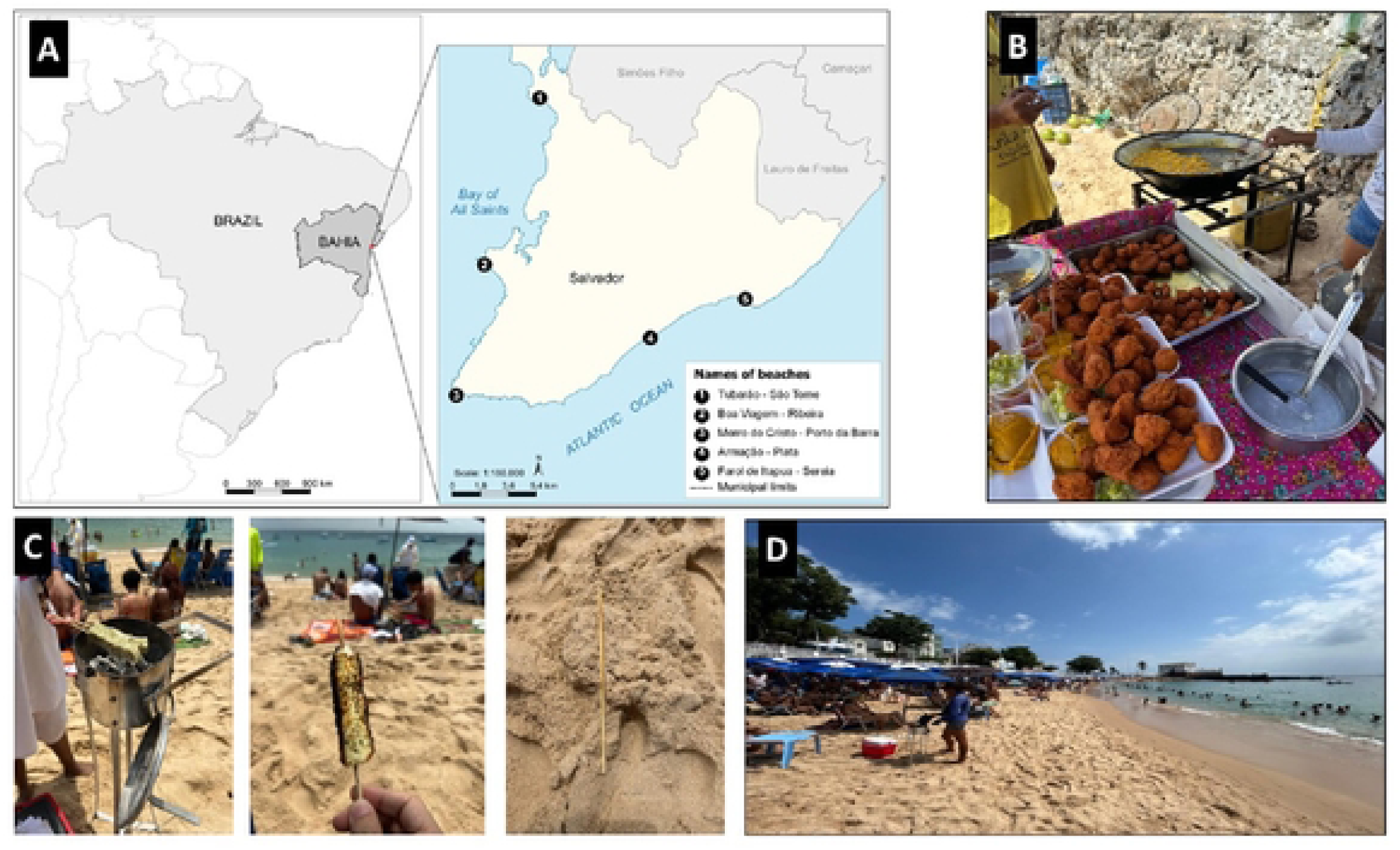
A) Map showing the location of the beaches in Salvador, Bahia, Brazil where data was collected; B) Typical informal sale of acarejá (bean cake) fried in hot palm oil at the beach; c) Grilling of coalho cheese on charcoal, with skewer left on the beach shore; D) A barefooted water vendor at Porto da Barra Beach, highlighting the risk of stepping on discarded skewers.

### Sampling process

To calculate the sample size, a total of 8,355 workers was used as the reference population, based on the number of street vendors registered to work at the 2023 Carnival in Salvador-a major festival in the city(14). Using these details, the sample size was calculated with Epi Info^TM^ software from the Centers for Disease Control and Prevention(15), considering the following assumptions: a 30% prevalence for WAs(5), a 95% confidence level and a 5% margin of error. This calculation distributed across five study areas, resulted in a minimum required sample size of 315 participants, with at least 63 from each beach area.

Inclusion criteria required workers to be at least ≥ 14 years old and have engaged in economic activities on the beaches for a minimum of three months, working at least once a week. Exclusion criteria included severe speech or hearing impairments, inability to communicate in Portuguese, and visibly being under the influence of alcohol or other psychoactive substances.

Participants were selected using two approaches. For mobile workers conducting economic activities along the beach, interviewers stationed themselves at central fixed points, visually identifying and counting workers as they passed. Every third eligible worker was approached and invited to participate in the study. For workers in fixed locations, interviewers walked the length of the beach, identifying workstations from left to right, facing away from the sea. Every third workstation was visited, and all eligible workers were invited to participate in the study.

### Data collection

This took place on Mondays, Wednesdays and Fridays, from 8 am to 12 pm, from November 6, 2023 to March 8, 2024. A team of 6-8 trained interviewers administered previously standardized and validated questionnaires, ensuring adherence to ethical standards. The data collection process involved two preliminary stages before the main study: a pre-pilot phase, in which the procedures were tested among the interviewers, and a pilot study conducted among six beach workers (not included in the final sample) between August and October 2023. The questionnaire encompassed sections on sociodemographic and socioeconomic information, occupational details, comorbidities, mental health, quality of life, work environment and processes, and access to health and social assistance services. Interviews were conducted using Research Electronic Data Capture-REDCap(16). Each interviewer wore a project-customised uniform for easy identification and conducted interviews in a location conducive to participants.

The final database was securely stored on a private server managed by the Institute of Collective Health (ISC), Federal University of Bahia (UFBA), with access restricted to the research coordinator. All procedures adhered to the guidelines of Brazil’s General Law on the Protection of Personal Data(17), ensuring the privacy and security of collected information.

### Sociodemographic and socioeconomic variables

To characterize the workers, the questionnaire focused on the following variables: sex (female, male); age (categorized as ≥60, 50-59, 40-49, 30-39, ≤29); race/ethnicity (classified as Black, Brown, White, Yellow/Indigenous); marital status (married/stable union, single, separated/widowed); education level (classified as primary education incomplete, primary education completed, secondary education incomplete, secondary education completed, tertiary education completed or incomplete); and persons with disability (no, yes).

### Occupational profile

To characterize the occupational profile, the following questions were asked: whether the participant’s income is exclusively from beach work (no, yes); average daily income on weekdays (classified into quartiles in USD: >41, 25-41, 14-24, <14) and on weekends (same quartiles); main economic activity on the beach (with 21 options, later grouped into four most frequent categories: food preparation and/or sale-stationary, waiter, sales and service provider and others); working method (stationary-sitting/standing, walking/moving, mixed); current employment condition on the beach (permanent, temporary, emergency due to recent job loss); current contribution to the National Institute of Social Security (INSS) (no, yes); daily working hours on the beach (classified into quartiles: ≤8, 9-10, 11-12, ≥13hrs); weekly working days (classified into tertiles: ≤5, 6, 7days); total time spent working on the beach in months (classified into quartiles: ≤24, 25-96, 97-240, ≥241); whether they carry weights while working (no, yes); use of personal protective equipment (PPE) (no, yes); and whether they have received relevant work-related training (no, yes).

### Work-related injuries or illnesses

All workers, regardless of whether they had reported a recent WA, were asked if they had experienced any workplace-related injuries and illnesses during their work in the past 12 months, specifically: exposure to biological material (e.g. bodily fluids, contaminated needles etc.), incidents involving venomous animals (e.g. snake bites) or cases of exogenous poisoning (e.g. contact of cleaning products with the skin, inhalation or ingestion). Also, workers were asked if a medical or health professional had diagnosed them with Noise-Induced Hearing Loss (NIHL), Repetitive Strain Injury (RSI)/Workplace-Related Musculoskeletal Disorders (WMSD) (response options: no, yes).

### Outcome variable

Information on WA history was gathered using the primary questions: “In the past 12 months, have you experienced any accidents at work or on your way to work?” with a binary answer (no, yes). For those who responded affirmatively, additional questions were asked: How many WAs have occurred in the past 12 months? (once, twice or more, often); for the most serious WAs, how did it occur/type of accident? (cut, puncture, burn, fall, others); what was the main part of the body affected? (head, upper limbs, torso, lower limbs); type of health damage or sequelae suffered due to the WA? (none, multiple, scars); did the WA cause you to miss work? (no, yes); and did you require rehabilitation or professional treatment? (no, yes).

### Data analysis

The sociodemographic and occupational characteristics, along with workplace-related injuries and illnesses in the past 12 months, were presented in absolute and relative values. The frequencies of the outcome (WA) among categories of independent variables were summarized using prevalence measures (Tables 1, 2 and 4). The Chi-square test for heterogeneity was used to assess observed versus expected variability within the sample, with the probability value (p-value) as a criterion for variable selection in the final model, all variables with p-value ≤0.20 were included. Crude prevalence ratios (CPR) and adjusted (aPR), with respective 95% confidence intervals, were estimated using a Poisson regression model with robust variance. In the final model (Table 3), a significance level of 5% (p≤0.05) was used to determine associations between WA history and sociodemographic and occupational variables. Data processing and statistical analyses were performed in R software, version 4.3.2(18). All datasets and codes used during this study are available in Zenodo under a Creative Commons license, accessible through(19).

**Table 1-.**
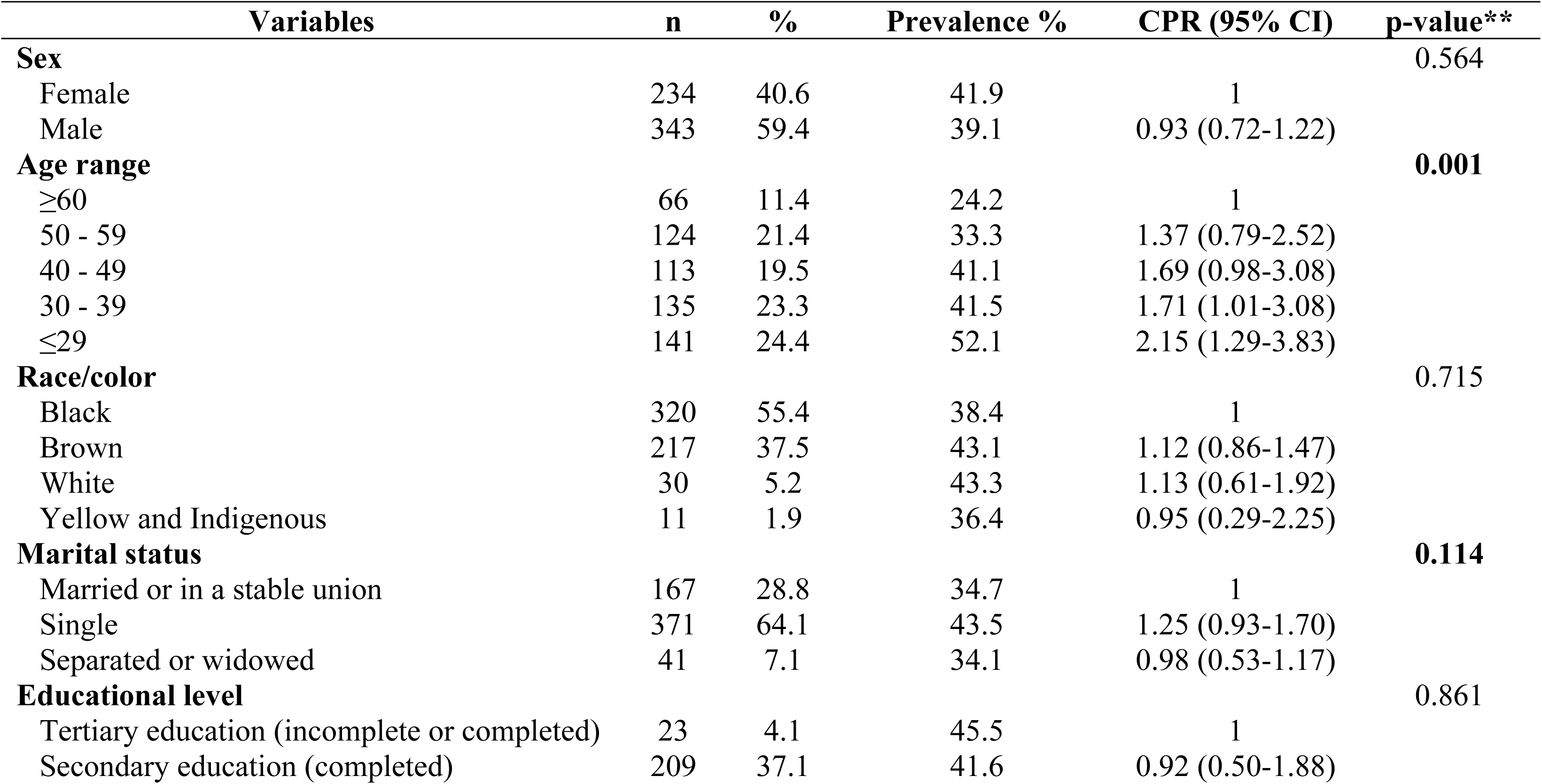
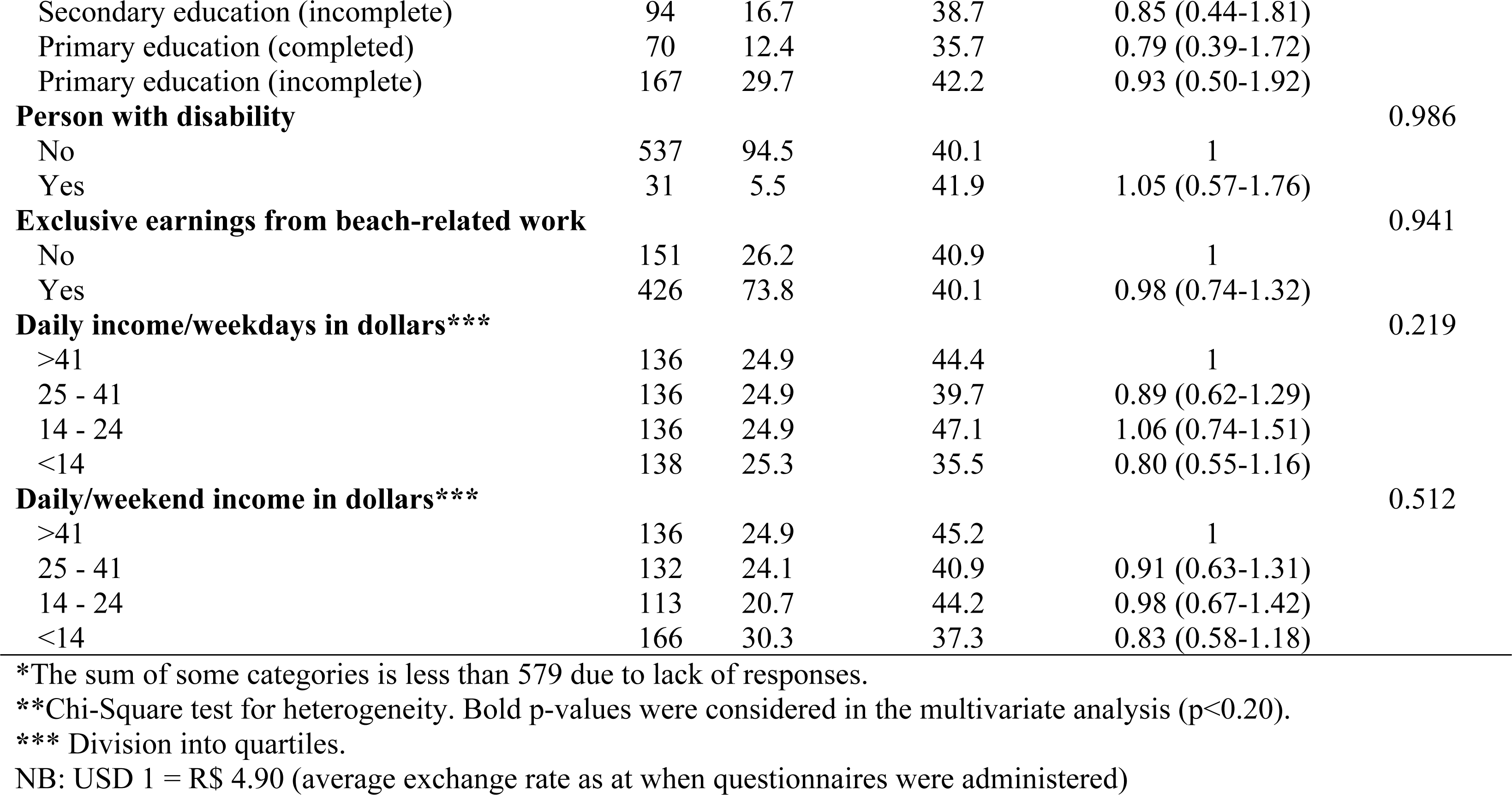
Sociodemographic and socioeconomic profile, prevalence and Crude Prevalence Ratio (CPR) of workplace accidents among urban beach workers in Salvador, 2023-2024. (n= 579)*

**Table 2-.**
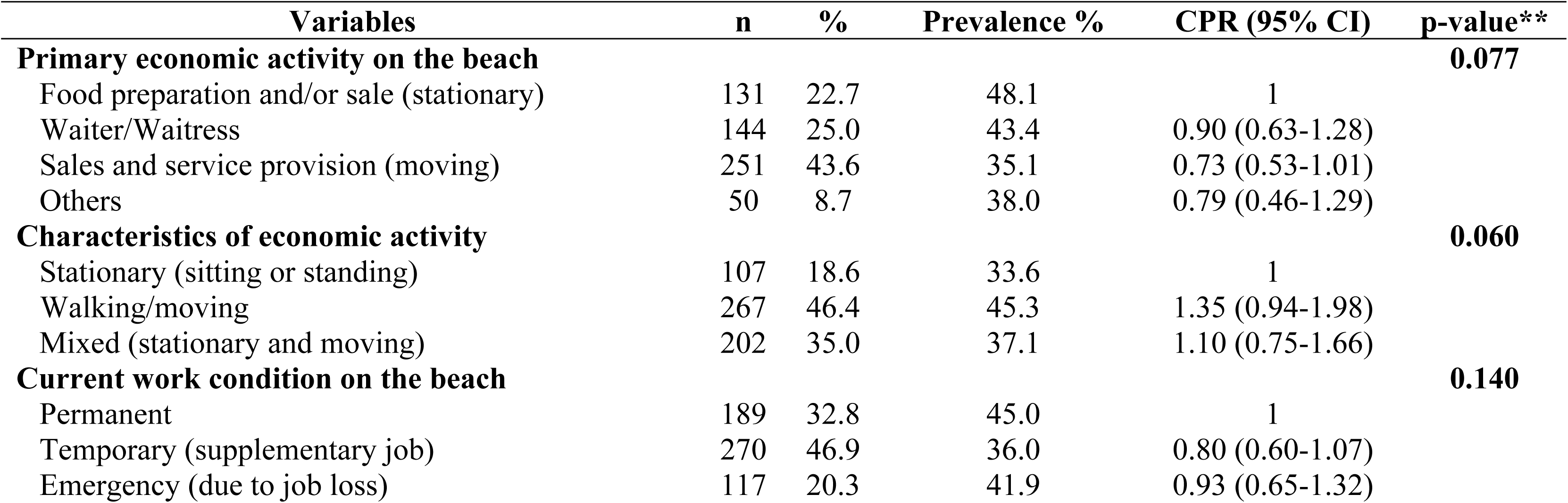
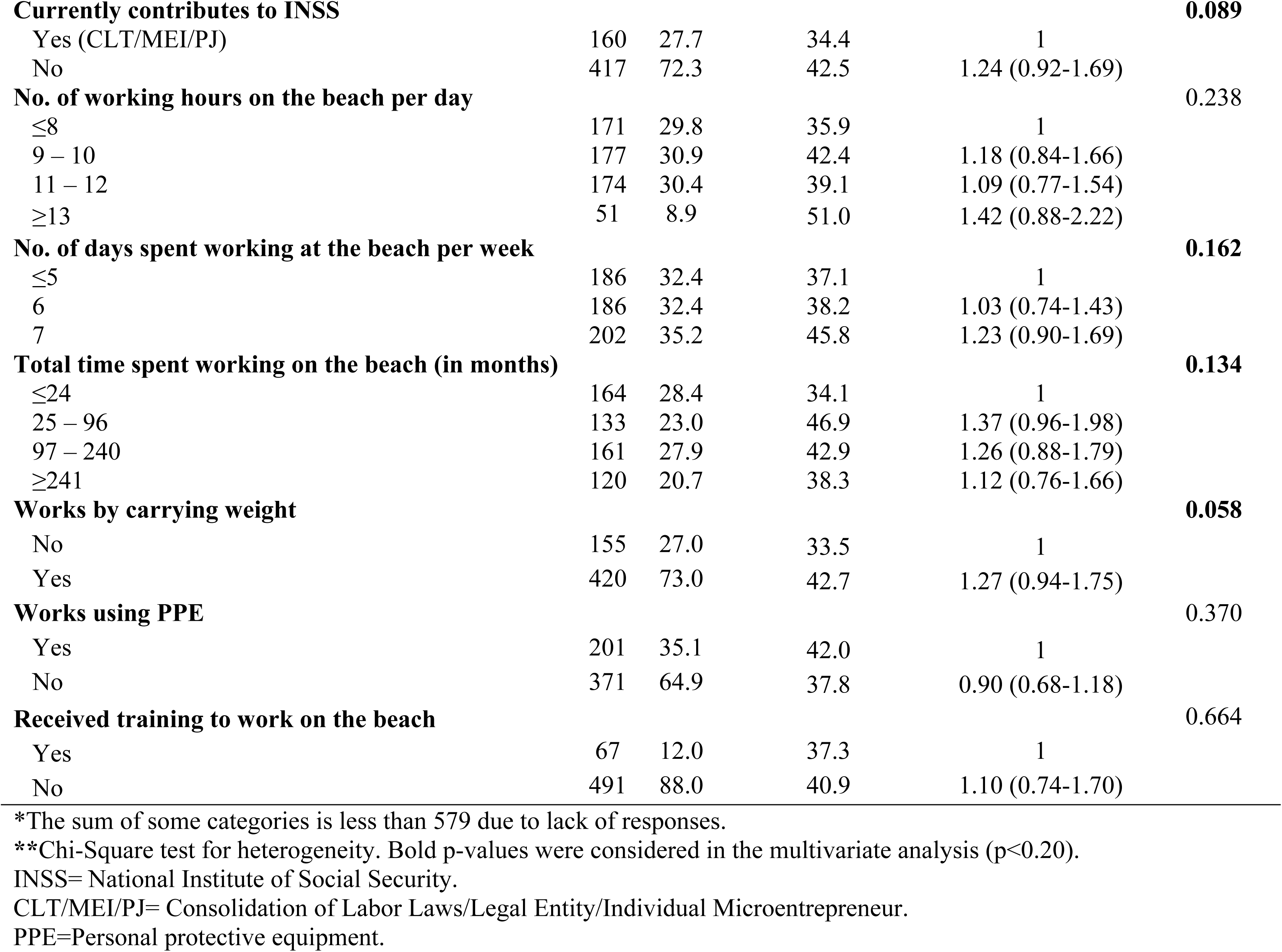
Occupational profile and work environment, prevalence and Crude Prevalence Ratio (CPR) of workplace accidents among urban beach workers in Salvador, 2023-2024. (n= 579)*

**Table 3-.** Final multivariate analysis model of adjusted Prevalence Ratio (aPR) for workplace accidents and associated factors among urban beach workers in Salvador, 2023-2024. (n=579)*

| Variables from the final model | aPR (95 % CI)** |
| --- | --- |
| **Age range** |  |
| ≥60 | 1 |
| 50 - 59 | 1.41 (0.79-2.62) |
| 40 - 49 | 1.77 (0.98-3.34) |
| 30 - 39 | 1.79 (0.98-3.45) |
| ≤29 | **2.59 (1.38-5.08)** |
| **Marital status** |  |
| Married or in a stable union | 1 |
| Single | 1.16 (0.85-1.60) |
| Separated or widowed | 1.20 (0.64-2.13) |
| **Primary economic activity on the beach** |  |
| Food preparation and/or sale (stationary) | 1 |
| Waiter/Waitress | 0.83 (0.57-1.22) |
| Sales and service provision (moving) | 0.78 (0.56-1.09) |
| Others | 1.01 (0.58-1.70) |
| **Characteristics of economic activity** |  |
| Stationary (sitting or standing) | 1 |
| Walking/moving | 1.19 (0.81-1.78) |
| Mixed (stationary and moving) | 0.98 (0.66-1.50) |
| **Current work condition on the beach** |  |
| Permanent | 1 |
| Temporary (supplementary job) | 0.85 (0.61-1.18) |
| Emergency (due to job loss) | 1.01 (0.68-1.50) |
| **Currently contributes to INSS** |  |
| Yes (CLT/MEI/PJ) | 1 |
| No | 1.12 (0.82-1.56) |
| **No. of working days on the beach per week** |  |
| ≤5 | 1 |
| 6 | 1.03 (0.73-1.46) |
| 7 | 1.20 (0.86-1.67) |
| **Total time spent working on the beach (in months)\*\*\*** |  |
| ≤24 | 1 |
| 25 - 96 | 1.35 (0.93-1.95) |
| 97 - 240 | 1.44 (0.97-2.14) |
| ≥241 | 1.49 (0.89-2.50) |
| **Works by carrying weight** |  |
| No | 1 |
| Yes | 1.22 (0.90-1.70) |

\*The sum of some categories is less than 579 due to lack of responses.
\*\*Prevalence Ratio values with respective 95% Confidence Interval in bold were statistically significant in the final model.
\*\*\*Division into quartiles.
INSS= National Institute of Social Security.
CLT/MEI/PJ= Consolidation of Labor Laws/Legal Entity/Individual Microentrepreneur.

### Ethical statement

The research project received approval from a Research Ethics Committee in line with CNS Resolution(20), of the National Health Council, under CAAE: 68859623.0.0000.5030 and opinion No. 6,501,806. All participants provided informed consent, agreeing to participate in the study and permitting the anonymous publication of the results, after reading and signing the Free and Informed Consent Form (FICF). For participants under 18 years of age, both the FICF and authorization form signed by a legal guardian were required.

## RESULTS

Of the 579 workers interviewed, 343 (59.4%) were male. Approximately 25% were young adults aged 29 years or younger, while only 11.4% were 60 years or older. Regarding race/color, 55.4% identified as Black and 37.5% as Brown, comprising 92.9% of participants. Most respondents (64.1%) reported being single. Regarding education, 58.8% had not completed high school, while just 37.1% had finished high school. People with disabilities represented 5.5% of the sample. In terms of occupational characteristics, 426 (73.8%) indicated that their primary income is derived from beach-related activities, with average daily income often below USD 24.00 on weekdays, and reaching up to USD 40.00 on weekends (Table 1).

The main economic activities on the beaches were vending and service provision (43.6%) and waitering (25%). These occupations influenced work patterns, with 46.4% of workers being mobile, frequently moving as they worked, while only 18.6% performed stationary tasks, either standing or seated. Temporary or emergency beach work was reported by 67.2% of participants, and 72.3% did not contribute to the INSS, highlighting the informal nature of their labor. Also, 70.2% worked over 8 hours/day, and 35.2% worked uninterruptedly throughout the week without days off. Notably, 73% reported carrying heavy loads during work, 64.9% indicated they did not use PPE, and 88% had never received any relevant job-related training from the local government or workers organizations (Table 2).

Among the 579 workers interviewed, 40.3% reported experiencing WAs within the past 12 months. WA prevalence was highest among female workers (41.9%), workers aged 29 or younger (52.1%), self-declared White workers (43.3%), single workers (43.5%) and those with higher education levels (45.5%). Additionally, WAs were more common among workers with disabilities (41.9%). In terms of occupational characteristics, WAs were most prevalent among those involved in food preparation or sales (48.1%), mobile workers (45.3%), those working permanently on the beaches (45%) and informal workers (42.5%). Higher WA rates were also observed among respondents working ≥13 hours per day (51%), those working 7 days a week (45.8%), workers carrying heavy loads (42.7%) and those lacking relevant job-related training (40.9%) (Tables 1 and 2).

The final multivariate model with adjusted prevalence ratio (aPR) for WAs in the past 12 months and associated factors among beach workers, is described in Table 3. Overall, a trend of increased WA probability was observed with decreasing age. Workers aged ≤29 years were significantly more likely to experience WAs, i.e. 2.59 times higher compared to workers aged ≥60 years (95%CI 1.38-5.08). Although other variables were not statistically significant in the final model, higher probabilities of WAs were observed among separated/widowed workers (PR: 1.20), those engaging in mobile economic activities (PR: 1.19), informal workers (PR: 1.12), workers with longer durations of beach-related economic activities (PR: 1.49) and those who carried heavy loads (PR: 1.22) (Table 3).

The profile of workplace-related accidents or injuries in the past 12 months, by sex, among workers on urban beaches in Salvador is described in Table 4. Among the 232 workers (40.3%) who reported experiencing WAs in the past 12 months, 42.2% experienced WAs more frequently. The most common types of accidents were punctures (42.7%) and cuts (28.4%), primarily affecting the lower limbs (70.6%). Regarding incidence outcomes, 33.7% of workers reported no lasting effects, while 54.7% reported scars from the incidents. Additionally, 27.4% required temporary leave from work, and 8.6% required rehabilitation or other treatment. When stratified by sex, frequent accidents were more prevalent among females than males (52% vs. 35.4%), with burns occurring five times more often among females than males (22.4% vs. 4.5%).

**Table 4-.** Profile of workplace-related injuries and illnesses by gender among urban beach workers in Salvador, 2023-2024. (n=579)*

| Variables | n | % | Prevalence n (%) |  | p-value** |
| --- | --- | --- | --- | --- | --- |
|  |  |  | Female | Male |  |
| **Workplace accident/route ≤12 months** |  |  |  |  | 0.564 |
| No | 344 | 59.7 | 136 (58.1) | 207 (60.9) |  |
| Yes | 232 | 40.3 | 98 (41.9) | 133 (39.1) |  |
| **Only workers with a history of WA (n=232)** |  |  |  |  |  |
| **Frequency of accident** |  |  |  |  | **0.029** |
| Only once | 64 | 27.6 | 20 (20.4) | 43 (32.3) |  |
| Twice or more | 70 | 30.2 | 27 (27.6) | 43 (32.3) |  |
| Often | 98 | 42.2 | 51 (52.0) | 47 (35.4) |  |
| **Type of accident** |  |  |  |  | **<0.001** |
| Cut | 66 | 28.4 | 21 (21.4) | 45 (33.8) |  |
| Punctures | 99 | 42.7 | 44 (44.9) | 54 (40.6) |  |
| Burn | 28 | 12.1 | 22 (22.4) | 6 (4.5) |  |
| Fall | 11 | 4.7 | 1 (1.0) | 10 (7.6) |  |
| Others (shock, drowning, being run over, etc.) | 28 | 12.1 | 10 (10.2) | 18 (13.5) |  |
| **Affected body part** |  |  |  |  | **0.042** |
| Head | 8 | 3.5 | 2 (2.0) | 6 (4.5) |  |
| Upper limbs | 55 | 23.8 | 21 (21.5) | 34 (25.8) |  |
| Torso | 5 | 2.1 | 5 (5.1) | 0 (0.0) |  |
| Lower limbs | 163 | 70.6 | 70 (71.4) | 92 (69.7) |  |
| **Effect of OA** |  |  |  |  | 0.225 |
| None | 78 | 33.7 | 27 (27.6) | 51 (38.3) |  |
| Multiple | 27 | 11.6 | 13 (13.3) | 14 (10.5) |  |
| Scars | 127 | 54.7 | 58 (59.2) | 68 (51.1) |  |
| **Temporarily required time away from work** |  |  |  |  | 0.818 |
| No | 167 | 72.6 | 72 (74.2) | 95 (72.0) |  |
| Yes | 63 | 27.4 | 25 (25.8) | 37 (28.0) |  |
| **Needed rehabilitation/treatment** |  |  |  |  | 0.994 |
| No | 212 | 91.4 | 89 (90.8) | 122 (91.7) |  |
| Yes | 20 | 8.6 | 9 (9.2) | 11 (8.3) |  |
| **workplace-related injuries and illnesses (n=576)** |  |  |  |  |  |
| **Exposure to biological material** |  |  |  |  | 0.999 |
| No | 451 | 78.3 | 183 (78.2) | 266 (78.2) |  |
| Yes | 125 | 21.7 | 51 (21.8) | 74 (21.8) |  |
| **Accident with venomous animal** |  |  |  |  | 0.450 |
| No | 548 | 95.1 | 225 (96.2) | 321 (94.4) |  |
| Yes | 28 | 4.9 | 9 (3.8) | 19 (5.6) |  |
| **Exogenous intoxication** |  |  |  |  | 0.818 |
| No | 542 | 94.1 | 219 (93.6) | 321 (94.4) |  |
| Yes | 34 | 5.9 | 15 (6.4) | 19 (5.6) |  |
| **Diagnosis of NIHL** |  |  |  |  | **0.018** |
| No | 550 | 95.6 | 229 (98.3) | 319 (93.8) |  |
| Yes | 25 | 4.4 | 4 (1.7) | 21 (6.2) |  |
| **Diagnosis of RSI/WMSD** |  |  |  |  | **<0.001** |
| No | 471 | 81.9 | 175 (75.1) | 294 (86.5) |  |
| Yes | 104 | 18.1 | 58 (24.9) | 46 (13.5) |  |
| **Diagnosis of skin diseases/infections** |  |  |  |  | 0.193 |
| No | 427 | 74.5 | 167 (71.4) | 258 (76.6) |  |
| Yes | 146 | 25.5 | 67 (28.6) | 79 (23.4) |  |

\*The sum of some categories is less than 579 due to lack of responses.
\*\*Chi-Square test for heterogeneity. Statistically significant P-values (p<0.05) are highlighted in bold.
NIHL=Noise-Induced Hearing Loss.
RSI/DORT= Repetitive Strain Injuries and Workplace-Related Musculoskeletal Disorders.

All interviewees, regardless of having experienced a WA in the past 12 months, were asked about other workplace-related injuries. Overall, 21.7% of the workers reported exposure to biological material, 4.9% experienced accidents involving venomous animals, and 5.9% reported cases of exogenous poisoning. Diagnosed conditions included NIHL in 4.4% of workers, RSI and WMSDs in 18.1%, and skin infections in 25%. Sex-based comparison revealed that NIHL was higher among males than females (6.2% vs. 1.7%), while RSI/WMSDs were more frequently reported by females than males (24.9% vs. 13.5%).

## DISCUSSION

This study aimed to characterize the sociodemographic and occupational profile and to assess the prevalence of WAs among beach workers in Salvador, Bahia, Brazil, over the past 12 months. Findings identified a 40.3% prevalence of WAs, with higher rates among younger workers, particularly those aged ≤29 years, where one in every two respondents reported a WA. Most accidents affected the lower limbs, with punctures being the most frequent injury type. Following a WA, approximately 25% of workers required temporary leave, potentially impacting both personal and family economic stability, given that 72% of workers were informally employed and lacked access to social security benefits. These findings provide new insights into a segment of the workforce in Brazil’s coastal cities, highlighting likely deficiencies in workplace safety and a gap in professional training policies for most informal employees.

Salvador’s economy is driven by commerce, services, and tourism. Bordered by the Atlantic Ocean and the Bay of All Saints (Figure 1A), the city features extensive beach areas where numerous workers engage in various economic activities (Figure 1D). Here, workers reported an average monthly income of R$2,060.00/US$375.00 (data not shown in tables), which is less than half the average salary of formal workers in the city (R$4,200.00/US$760.00 in 2022)(13). On average, respondents worked 10 hours a day and 35% reported working without any days off, maintaining continuous economic activities throughout the week. High levels of informality, limited education, and the fact that 67% of the workers viewed their beach-related activities as temporary or emergency work likely contribute to their lower incomes and extended work hours.

Overall, 40.3% of workers reported experiencing at least one WA in the past 12 months. Given the limited availability of studies on similar populations, our findings were compared with data from workers in other informal sectors potentially exposed to high WA prevalence. For instance, a systematic review and meta-analysis reported a slightly higher WA prevalence of 46.8% among construction workers in Ethiopia(21), slightly exceeding the 40.3% observed in this study. Conversely, significantly lower prevalence rates were reported among agricultural workers-10%(22) and motorcyclists delivering goods-25%(23). A study on informal workers in the commerce sector of Jequié, southeastern-Bahia, predominantly food and beverage vendors, reported a 12-month WAs prevalence of 32%(5). The elevated prevalence of WAs in this study likely stems from multiple factors. Particularly, the high levels of informality and precarious working conditions are probable key contributors. Also, the demanding workloads, with many workers exceeding 8-hour shifts and working seven days a week may contribute to physical and mental fatigue, consequently reducing workers attention to work practices and increasing their risk to WAs(24,25).

The likelihood of experiencing a WA was higher among younger workers, with those aged ≤29 years being 2.5 times more likely to report WAs compared to workers aged ≥60 years. This pattern aligns with another finding conducted in Salvador between 2001 and 2004, which similarly reported a higher concentration of WAs among workers aged 18 to 30 years(26). Another study on informal workers in Bahia’s retail sector also observed high WA risk with an OR of 4.62 (95% CI: 1.82-11.73) for WA among workers ≤30 years compared to those aged 30 to 59 years(5). This trend may stem from the greater physical vigor of young workers, enabling them to engage in more frequent and intensive economic activities, thereby increasing their exposure to occupational hazards. Similarly, the lack of PPE use and limited work experience or training among younger workers could further heighten their vulnerability to WAs compared to older and more experienced counterparts.

Although the association between WAs by gender was not statistically significant, a slightly higher prevalence of WA was observed among female workers compared to males (41.9% vs. 39.1%). This is similar to the findings from a population-based study conducted in the same municipality, which reported a greater vulnerability to WAs among females compared to males (64% vs. 36%)(26). Although the study by Santana et al.(26) did not specify the occupations of the injured participants, several characteristics of the participants, such as high levels of informality (54.3%), low education levels(51.4%) and low economic status (55.1%) are comparable to those observed in this study which could be predisposing participants to WAs.

When stratified by sex, burns emerged as the second most reported cause of WAs among female workers (22.4%), occurring five times more frequently than among males (4.5%). This disparity may be explained by the greater involvement of females in preparing hot and fried foods, particularly acarajé (beans cake)-a popular delicacy in Salvador made from beans and fried in hot palm oil, typically prepared in large pans under improvised conditions (Figure 1B). Since acarajé preparation is predominantly carried out by women, it may increase their exposure to burn-related risks during work.

The high proportion of puncture-related injuries and lower limb accidents reflects the nature of predominant activities among vendors and waiters, who are constantly on the move. A key factor contributing to this pattern of WAs is the rampant consumption of coalho cheese, another popular delicacy grilled and sold on wooden skewers across many beaches in Salvador (Figure 1C). These sharp skewers are often discarded carelessly, posing a considerable risk of puncture injuries to workers navigating the beach during their daily activities.

Expectedly, skin infections were the most prevalent workplace-related injuries and illnesses, affecting 25.5% of workers. Similarly, a cross-sectional study conducted in northeastern Brazil among beach workers in Natal, the capital of Rio Grande do Norte, found a prevalence of 27.1% of lip lesions(12). While the current study did not specify the type or location of infections, the high prevalence found may be the effect of chronic exposure to solar radiation, frequent contact with sand and saltwater, and the significant proportion of workers (64.9%) not using PPEs.

Although this study observed a high prevalence of RSI/WMSDs (18.1%), the observed rate is notably lower than findings in other occupational settings. For example, systematic reviews and meta-analyses by Tolera et al.(27) reported a prevalence of 40% among urban cleaners, while Khoshakhlagh et al.(28) found 41% among firefighters, and Wang et al.(29) documented 79% among nurses. In Brazil, a report by the Ministry of Health (30), using national notification data, estimated 9.6 cases of RSI/WMSDs per 100,000 workers in 2016, reflecting a 170% increase compared to 2007. As in the present study, the data from the Ministry of Health showed a higher proportion among females. Due to the great physical effort required to perform activities on beaches — with 73% of respondents reporting carrying weight during work — an even higher prevalence of RSI/WMSDs than that found would be expected. It is worth mentioning that during the interviews, workers were specifically asked if they had received a diagnosis from a doctor or health professional within the past 12 months. This may have led to underreporting, as some workers could have RSI/WMSD but lacked access to health services for proper diagnosis or chose not to be diagnosed to avoid interruptions in their economic activities.

The observed 4.4% prevalence of NIHL in this study, with rates 3.6 times higher among males, may be associated with persistent exposure to loud noises in beach environments. These often stem from multiple individual speakers used by bathers, combined with noise from nearby commercial establishments, a common occurrence in Salvador. Such exposure could subject workers to constant noise levels potentially exceeding the WHO-recommended limit of 85 dB(A)(31). While this study did not include measurements of local noise levels, no workers were observed using any form of sound protection device. Future studies to measure noise levels on beaches, in addition to audiometric tests on workers, are necessary.

The high prevalence of workers reporting exposure to biological material (21.7%) is unexpected, as such occupational exposure is typically associated with healthcare workers who frequently handle body fluids and physical agents(32,33). While this finding cannot be entirely dismissed, it raises the possibility of misinterpretation by respondents. Workers who suffered puncture-related WAs, particularly those involving skewers, cans or glass may have misinterpreted them as WAs involving exposure to biological material. This potential misclassification underscores the need for future research to clarify the nature and context of these exposures, ensuring a more accurate understanding and interventions for this scenario.

Although Salvador has a high proportion of informal workers (42%)(13), this study observed that 72% of beach workers belong to this condition. However, this proportion is lower than the 94.5% informality reported among 362 beach workers in a 2010 survey conducted in Natal, Brazil(12). The proportion of informal beach workers remains one of the highest and surpasses the national average, which stood at 40% of the 101 million employed Brazilians in the first half of 2024(13). Globally, around 2 billion workers were in informal employment over the past decade, with rates ranging from a maximum of 20% in North America and Western Europe, including countries like Spain, England and Italy, to over 90% in some African nations such as Angola, Nigeria, Ghana and the Republic of Congo. Across these regions, informality is disproportionately higher among women, non-white individuals, those with lower education levels, and workers in occupations that require minimal technical expertise-demographics that partly align with the profile of beach workers(13, 34). As a result of informality, workers experience significant disadvantages, particularly the absence of social protections, such as sick leave, accident compensation, maternity leave, and retirement benefits. Consequently, these individuals are often compelled to continue working despite unfavorable health conditions or precarious work environments as reflected in our findings here.

Another plausible explanation for the high prevalence of workplace-related injuries and illnesses observed in this study could be the lack of relevant job-related training among the surveyed population. Only 12% of respondents reported having received any form of training, and nearly 73% indicated they had no access to preparatory courses related to their activities(11). This gap may have been compounded by the absence of specialized training programs tailored to the needs of beach workers. An online review of training opportunities offered by Salvador’s local governments revealed only courses focused on tourism and entrepreneurship, with none particularly addressing beach-related activities(35). At the national level, the National Service for Commercial Training (SENAC) offers free courses in areas like languages, information technology, tourism and construction yet lacks specific programs targeting beach workers(36), highlighting the invisibility of this group by the policymakers.

The authors acknowledge certain limitations in the study’s design. For example, the sample size calculation relied on the 8,355 workers registered on the city hall website, a figure that primarily reflects those granted annual licenses to work during major events such as Carnival(14). However, given that 42% of Salvador’s 950,000 workers are informal, it is likely that many individuals engaging in economic activities on the beaches may not be officially registered. This could have resulted in an underrepresentation and to address this, the final population size was almost doubled, from the calculated minimum of 315 to 579 respondents. Also, the selection of beaches was determined empirically, based on the visual concentration of street vendors along Salvador’s 50-kilometers coastline. The five beaches cover 17 of Salvador’s 33 urban beaches (51.5%)(37), ensuring geographic representation of the city (Figure 1A).

Given the length of the questionnaire, which required an average of 30 minutes to complete, and the value of respondents’ time, interviews were exclusively conducted on weekdays between 8 am and 11 am, a period typically characterized by lower activity levels. To ensure a comfortable environment for both interviewers and respondents, the interviews were conducted in a designated space provided by the research team. This space included a tent for shade and water supply, offering a basic level of comfort during the data collection process.

Given the characteristics of the study and investigated population, the healthy worker effect warrants consideration. The high proportion of informal workers in the study population indicates that many continue their economic activities despite adverse physical and/or mental conditions. However, it is possible that the sample does not entirely represent the broader beach workforce, potentially excluding individuals with severe health conditions that prevented them from working at the time of the interviews. As a result, the actual prevalence of workplace-related injuries and illnesses in this population may be underestimated.

The cross-sectional design of the study restricted the analysis to period prevalence, focusing on the past 12 months. This design does not permit longitudinal monitoring of the potential causes, such as sociodemographic and occupational characteristics, and the outcome of interest (i.e. WAs). To address the potential for reverse causality, questions with temporal markers were incorporated into the data collection instrument

Despite the limitations highlighted, this study makes a significant contribution to the limited body of literature on the epidemiological, occupational and historical profile of WAs among beach workers. The sampling strategies, participant selection process, and data collection methodology employed can serve as a framework for replicating the study in other LMICs with similar ecological and sociodemographic characteristics. To ensure data reliability, regular training and retraining sessions were conducted for the research team before data collection. Additionally, data analyses stratified by interviewer were performed simultaneously to minimize interviewer bias. To reduce recall bias, questions concerning workplace-related injuries and illnesses were limited to the previous 12 months. Finally, the findings of this study should offer critical insights for managers and employers to enhance work environments and processes, fostering safer conditions for workers to conduct their daily activities on the beaches of Brazil and in other countries with similar settings.

## CONCLUSIONS

Beach workers in Salvador, Bahia, Brazil, reported a high prevalence of WAs over the past 12 months, with 40.3% experiencing at least one episode. Workers aged ≤29 years had a 2.59 times higher probability of WAs compared to those aged ≥60 years. Although not statistically significant, trends indicated a higher occurrence of WAs among workers without formal employment contracts, those working every day, those who regularly carried heavy loads, and those with longer tenures in the roles. Among workplace-related injuries and illnesses, skin infections (25.5%), exposure to biological material (21.7%), and RSI/WMSDs (18.1%) were notably prevalent. These findings suggest that future interventions should focus on addressing adverse working conditions, including high informality, long working hours, and a lack of training to reduce WAs and improve workers’ overall well-being.

## Data Availability

All datasets and codes used during this study are available in Zenodo under a Creative Commons license, accessible through https://doi.org/10.5281/zenodo.14175754

## Funding

This study was supported by the “Ministério Público do Trabalho - 5ª região, Salvador, Bahia, Brazil”.

## Competing Interests

The authors have declared that no competing interests exist.

## Notes

### Competing Interest Statement

The authors have declared no competing interest.

### Author Declarations

The research project received approval from the Research Ethics Committee (CEP) of the Instituto de Saúde Coletiva, Universidade Federal da Bahia (ISC/UFBA) in line with CNS Resolution(20), of the National Health Council, under CAAE: 68859623.0.0000.5030 and opinion No. 6,501,806. All participants provided informed consent, agreeing to participate in the study and permitting the anonymous publication of the results, after reading and signing the Free and Informed Consent Form (FICF). For participants under 18 years of age, both the FICF and authorization form signed by a legal guardian were required.

## REFERENCES

1. Descatha A, Sembajwe G, Baer M, Boccuni F, Di Tecco C, Duret C, et al. WHO/ILO work-related burden of disease and injury: Protocol for systematic reviews of exposure to long working hours and of the effect of exposure to long working hours on stroke. Environ Int. 2018 Oct;119:366–78.

2. Ministério da Saúde. Coleção Visat Volume 2 - Orientações técnicas para a vigilância epidemiológica de óbitos por causas externas relacionadas ao trabalho acidentes de trabalho. Available from: https://www.gov.br/saude/pt-br/centrais-de-conteudo/publicacoes/svsa/saude-do-trabalhador/colecao-visat-volume-2.pdf/view. 2024 [cited 2024 Nov 18]

3. Lu L, Huang H, Wei J, Xu J. Safety regulations and the uncertainty of work-related road accident loss: The triple identity of Chinese local governments under principal-agent framework. Risk Anal. 2020 Jun;40(6):1168–82.

4. García-Mainar I, Montuenga VM. Risk self-perception and occupational accidents. J Safety Res. 2024 Feb;88:135–44.

5. Rios MA, Nery AA, Rios PAA, Casotti CA, Cardoso JP. Fatores associados a acidentes de trabalho envolvendo trabalhadores informais do comércio. Cad Saude Publica. 2015 Jun;31(6):1199–212.

6. Takala J, Hämäläinen P, Sauni R, Nygård CH, Gagliardi D, Neupane S. Global-, regional- and country-level estimates of the work-related burden of diseases and accidents in 2019. Scand J Work Environ Health. 2024 Mar 1;50(2):73–82.

7. Ministério da Saúde, Informações de Saúde. Available from: http://tabnet.datasus.gov.br/cgi/tabcgi.exe?sinannet/cnv/acgrbr.def. [cited 2024 Nov 18]

8. Kyung M, Lee SJ, Dancu C, Hong O. Underreporting of workers’ injuries or illnesses and contributing factors: a systematic review. BMC Public Health. 2023 Mar 24;23(1):558.

9. Tucker S, Diekrager D, Turner N, Kelloway EK. Work-related injury underreporting among young workers: prevalence, gender differences, and explanations for underreporting. J Safety Res. 2014 Sep;50:67–73.

10. Tabela 4093: Pessoas de 14 anos ou mais de idade, total, na força de trabalho, ocupadas, desocupadas, fora da força de trabalho, em situação de informalidade e respectivas taxas e níveis, por sexo. Available from: https://sidra.ibge.gov.br/tabela/4093. [cited 2024 Nov 18]

11. Xavier DG, Falcão JT, Lima CT. Caracterização da atividade laboral de trabalhadores informais em praia de Natal (RN) - Brasil. Cad Psicol Soc Trab. 2015;18.

12. Lucena EE de S, Costa DCB, Silveira ÉJD da, Lima KC de. Prevalência de lesões labiais em trabalhadores de praia e fatores associados. Rev Saude Publica. 2012 Dec;46(6):1051–7.

13. IBGE. Available from: https://cidades.ibge.gov.br/brasil/ba/salvador/panorama. [cited 2024 Nov 18]

14. Prefeitura divulga lista com 8.355 mil ambulantes aptos a trabalharem nas festas populares de Salvador – SEMOP. Available from: https://ordempublica.salvador.ba.gov.br/prefeitura-divulga-lista-com-8-355-mil-ambulantes-aptos-a-trabalharem-nas-festas-populares-de-salvador/. [cited 2024 Nov 18]

15. Epi InfoTM|CDC. (n.d.). Available from: https://www.cdc.gov/epiinfo/por/pt_index.html. 2024 [cited 2024 Nov 18]

16. REDCap. Available from: https://www.project-redcap.org/. [cited 2024 Nov 18]

17. L13709. (n.d.). Available from: https://www.planalto.gov.br/ccivil_03/_ato2015-2018/2018/lei/l13709.htm. [cited 2024 Nov 18]

18. R Core Team. R: A language and environment for statistical computing. R Foundation for Statistical Computing, Vienna, Austria. URL (2019) https://www.R-project.org

19. Cremonese C, Silva J dos S, Awoniyi MA. Code used to analyze data on work accidents among beach workers in Salvador, Bahia, Brazil, 2023-2024. Zenodo; 2024. Available from: 10.5281/zenodo.14175754

20. Conselho Nacional de Saúde. Conselho Nacional de Saúde. Available from: https://www.gov.br/conselho-nacional-de-saude/pt-br. [cited 2024 Nov 18]

21. Ashuro Z, Zele YT, Kabthymer RH, Diriba K, Tesfaw A, Alamneh AA. Prevalence of work-related injury and its determinants among construction workers in Ethiopia: A systematic review and meta-analysis. J Environ Public Health. 2021 Jul 24;2021:9954084.

22. Hunsucker S, Reed DB. Obesity and work-related injuries among farmers in Kentucky, Tennessee, and West Virginia. Workplace Health Saf. 2021 Dec;69(12):573–9.

23. Prakobkarn P, Luangwilai T, Prempree P, Kunno J. Prevalence of motorcycle accidents among food delivery drivers and its relation to knowledge, attitudes, and practices in urban areas in Bangkok, Thailand. PLoS One. 2024 May 23;19(5):e0303310.

24. WHO/ILO Joint Estimates of the Work-related Burden of Disease and Injury. Available from: https://www.who.int/teams/environment-climate-change-and-health/monitoring/who-ilo-joint-estimates. [cited 2024 Nov 18]

25. Gómez-García AR, Merino-Salazar P, Reiban G, Yela R. Jornadas laborales prolongadas y lesiones por accidentes de trabajo: estimaciones de la Primera Encuesta sobre Condiciones de Seguridad y Salud en el Trabajo en Ecuador. Arch Prevencion Riesgos Laborales. 2022;26:25–40.

26. Santana VS, Araújo GR de, Espírito-Santo JS do, Araújo-Filho JB de, Iriart J. A utilização de serviços de saúde por acidentados de trabalho. Rev Bras Saúde Ocup. 2007 Jun;32(115):135–44.

27. Tolera ST, Assefa N, Gobena T. Global prevalence of musculoskeletal disorders among sanitary workers: a systematic review and meta-analysis. Int J Occup Saf Ergon. 2024 Mar;30(1):238–51.

28. Khoshakhlagh AH, Yazdanirad S, Al Sulaie S, Mohammadian-Hafshejani A, Orr RM. The global prevalence of musculoskeletal disorders among firefighters: a systematic review and meta-analysis. Int J Occup Saf Ergon. 2024 Mar;30(1):272–91.

29. Wang K, Zeng X, Li J, Guo Y, Wang Z. The prevalence and risk factors of work-related musculoskeletal disorders among nurses in China: A systematic review and meta-analysis. Int J Nurs Stud. 2024 Sep;157(104826):104826.

30. Ministério da Saúde. Available from: http://editora.saude.gov.br. [cited 2024 Nov 18]

31. Functions S. World report on hearing [Internet]. World Health Organization; Available from: https://www.who.int/publications/i/item/world-report-on-hearing. 2021 [cited 2024 Nov 18].

32. Bertelli C, Martins BR, Reuter CP, Krug SB. Acidentes com material biológico: fatores associados ao não uso de equipamentos de proteção individual no Sul do Brasil. Cienc Amp Saude Coletiva Mar. 2023;28:789–801.

33. Reis LA, Gómez La-Rotta EI, Diniz PB, Aoki FH, Jorge J. Occupational Exposure to Potentially Infectious Biological Material Among Physicians, Dentists, and Nurses at a University. Saf Health Work. 2019 Dec;10(4):445–451. doi: 10.1016/j.shaw.2019.07.005.

34. International Labour Organization. A call for safer and healthier working environments. Available from: https://www.ilo.org/publications/call-safer-and-healthier-working-environments. 2023 [cited 2024 Nov 18].

35. Quali – Programa de Qualificação e Certificação das Empresas de Salvador [Internet]. Available from: https://quali.salvador.ba.gov.br/. [cited 2024 Nov 18].

36. Senac Cursos. Available from: https://cursos.ba.senac.br/. [cited 2024 Nov 18]

37. Instituto do Meio Ambiente e Recursos Hídricos (INEMA). Sistema deMonitoramento de Balneabilidade da Bahia. Salvador: INEMA. Disponível em: http://balneabilidade.inema.ba.gov.br/#resultado-pesquisa.

